# Third COVID-19 Vaccine Dose Boosts Neutralising Antibodies in Poor Responders

**DOI:** 10.1101/2021.11.30.21266716

**Authors:** Douglas F. Lake, Alexa J. Roeder, Maria J. Gonzalez-Moa, Megan Koehler, Erin Kaleta, Paniz Jasbi, John Vanderhoof, Davis McKechnie, Jack Forman, Baylee Edwards, Alim Seit-Nebi, Sergei Svarovsky

## Abstract

**Objective:** To determine if poor responders to COVID-19 RNA vaccines (<50% neutralisation) after two doses would remain poor responders, or if a third dose could elicit high levels of NAbs.

**Design:** Clinical follow-up study

**Setting:** Academic and medical institutions in USA

**Participants:** 269 healthy individuals ranging in age from 19 to 80 (Average age = 51; 165 females and 104 males) who received either BNT162b2 (Pfizer) or mRNA1273 (Moderna) vaccines.

**Main Outcome Measures:** NAb levels were measured: i) 2-4 weeks after a second vaccine dose, ii) 2-4 months after the second dose, iii) within 1-2 weeks prior to a third dose and iv) 2-4 weeks after a third RNA vaccine dose.

**Results:** In 269 study participants, percent neutralisation ranged from 0% to 99% 2-4 weeks after a second vaccine dose. The majority of vaccine recipients (154/269, 57%) demonstrated NAb levels at ≥75% 2-4 weeks after their second dose. Our study also revealed that 25% of vaccine recipients did not neutralise above 50% (Median neutralisation = 21%, titers <1:80) within a month after their second dose. We called these individuals “vaccine poor responders” (VPRs). Twenty-three VPRs ranging in age from 31 to 79 (10 males, 13 females, average age = 62.5) independently obtained a third dose of either BNT162b2 or mRNA-1273 vaccine 1-8 months (average = 5 months) after their second dose. Within a month after their third dose, poor responders showed an average 20-fold increase in NAb levels (range 46%-99%).

**Conclusions:** The results suggest that poor responders are not permanently poor responders; they can generate high NAb levels with an additional vaccine dose–independent of mRNA vaccine manufacturer. Previous reports indicate that NAb levels decline much more rapidly than clinical protection from hospitalisation and disease, but that does not account for vaccine recipients who never generated high levels of NAbs after two doses. It is possible that poor responders are a source of breakthrough infections. Although it is not known what levels of NAbs protect from infection or disease, many vaccine recipients in high-risk professions may wish to keep peripheral NAb levels high, limiting infection, asymptomatic viral replication, and potential transmission.

## INTRODUCTION

COVID-19 RNA vaccines prevent serious clinical disease requiring hospitalisation in ∼95% of vaccine recipients. This suggests that 5% of vaccinated individuals remain susceptible to infection and severe disease(1,2). If 300 million people receive two doses of the COVID-19 RNA vaccines, then approximately 15 million people may not be fully protected. Although T cells are important in anti-viral immunity, their activity is difficult to rapidly evaluate at scale. Furthermore, if T cells are engaged, the host is already infected. After natural infection with SARS-CoV-2 or vaccination against COVID-19, anti-viral antibodies are generated by the host. Except for antibodies that mediate antibody-dependent cellular cytotoxicity and complement dependent cytotoxicity, the only antibodies of primary importance are neutralising antibodies (NAbs). NAbs block the spike protein on SARS-CoV-2 from binding to the host cell receptor, angiotensin converting enzyme 2 (ACE2). In particular, the portion of the spike protein that binds to ACE2 is the receptor binding domain (RBD) (3,4) and there have been many reports of natural, vaccine-induced(5–8) and therapeutic antibodies(9) that neutralise the virus by binding to the RBD.

After 2 doses of either BNT162b2 or mRNA1273, antibodies to spike protein and neutralising antibodies have been quantified in vaccine recipients(1,2,10,11). Durability of those responses has been reported(12,13). Although it is not known what levels of NAbs provide protection against infection and potential disease, some individuals who are high-risk for COVID-19 or those that are caregivers for high-risk individuals may want to keep their NAb levels elevated to avoid asymptomatic infection and potential transmission to vulnerable populations. Since the vaccines do not elicit protective immunity in everyone, many vaccine recipients may want to know how well their vaccine induced protective antibodies and how long they circulate in peripheral blood. NAb levels have been modeled as correlates of protection from infection and/or disease(6). Here we report the results of a study in which NAb levels were measured in finger-stick whole blood from RNA vaccine recipients at 2-4 weeks and 2-4 months after their second dose, and then again pre and post 3^rd^ RNA vaccine dose.

In this report, we measured NAb levels using a semi-quantitative rapid test (14) in 269 healthy individuals ranging in age from 19 to 80 (Average age = 51; 165 females and 104 males) who received two doses of COVID-19 RNA vaccine. Twenty-three of those study participants received either three doses of BNT162b2 (Pfizer), 3 doses of mRNA-1273 (Moderna) or 2 doses of BNT162b2 followed by a third dose of mRNA-1273. Demographics of this cohort are shown in **Table S1**. The aim of this study was to determine if a 3^rd^ dose of vaccine in VPRs can generate strong neutralising antibodies.

## METHODS

Since performing neutralisation assays with authentic SARS-CoV-2 is time-consuming, expensive and requires high-containment facilities with specially trained laboratory personnel, we previously developed a rapid test that semi-quantitatively measures levels of neutralising antibodies in whole blood or serum. The rapid test utilises lateral flow technology and is based on the principle that NAbs of any isotype prevent the receptor binding domain (RBD) on spike protein from binding to ACE2 (**Figure 1**)(3,4). Interpretation if the test is counter-intuitive: the weaker the test line, the stronger the neutralising activity. Test and control line densities can be quantified with a lateral flow reader (iDetekt, Austin, TX) and recorded electronically.

**Figure 1.**
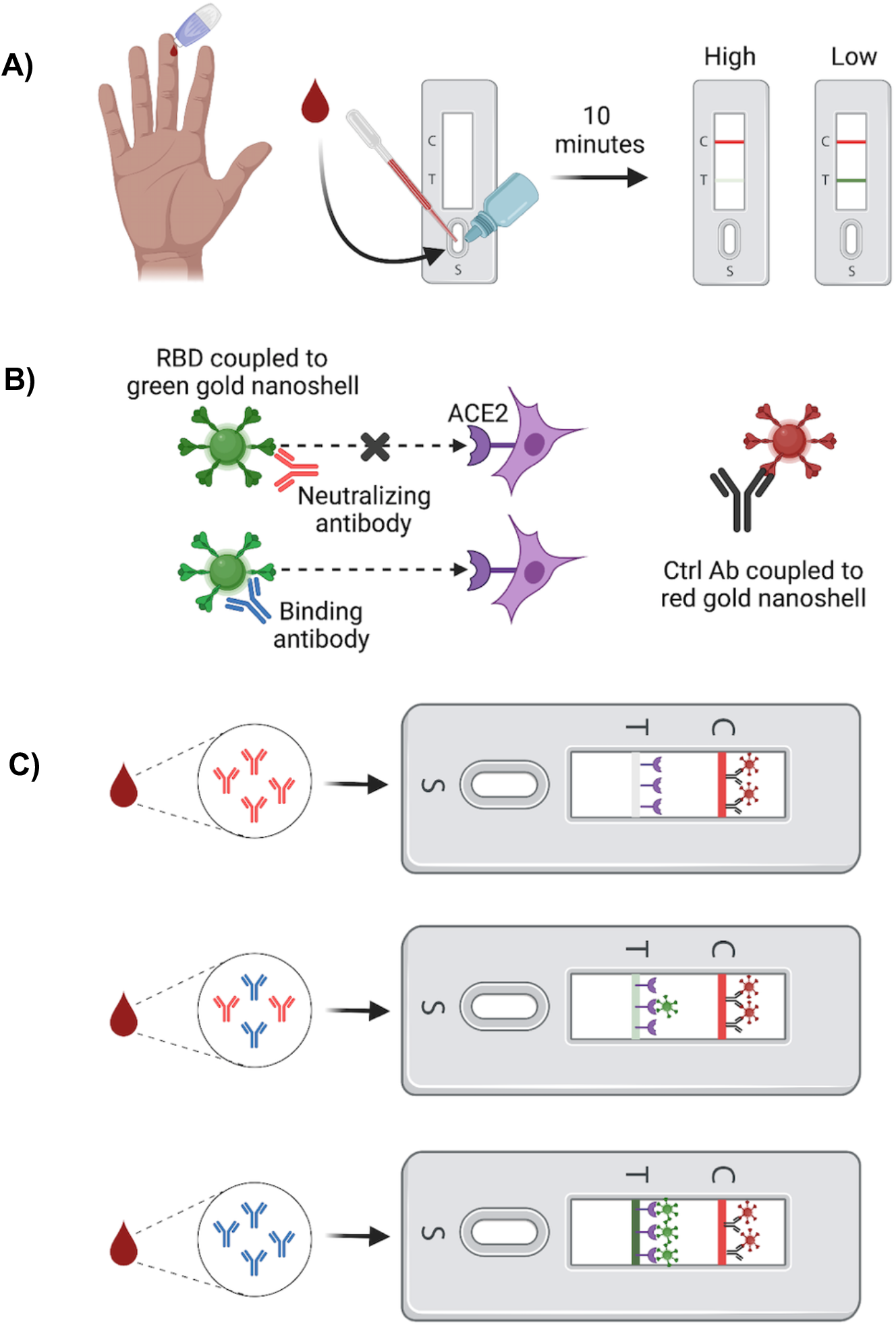
Rapid Test to Detect SARS-CoV-2 Neutralising Antibody. Lateral Flow Assay to Detect SARS-CoV-2 Neutralising Antibody. Interpretation of the rapid test is counter intuitive. **A)** Methodology overview. Fingerstick blood is transferred to the sample port and followed by two drops chase buffer. Ten minutes later results can be interpreted. Absent or faint test line indicates high NAb levels, while dark or intense test line indicates low/no NAbs. **B)** Mechanistic schematic. NAbs bind RBD coupled to a green gold nanoshell (GNS) and *prevent* the RBD/ACE2 interaction from occurring. Abs that bind RBD but do not neutralise *allow* the RBD/ACE2 interaction to occur, shown as increasingly dark signal as more RBD-GNS/ACE2 binds at the test line. **C)** Example test showing highly neutralising (top), moderately neutralising (middle), and poorly neutralising antibodies (bottom) within a blood sample. A monoclonal control antibody coupled to a red-GNS runs laterally with the sample/buffer mixture and binds at the control line, seen as a red line.

### Study Design and Participant Enrollment

Male and female adults ranging in age from 18-80 years old were recruited with informed consent to measure their NAb levels after vaccination with either BNT162b2 or mRNA1273. The study was approved by the internal review board at Arizona State University. In this cohort, no participant ever tested positive by PCR or was diagnosed with COVID-19 prior to the study. NAb levels were measured in all participants 2 to 4 weeks after a second dose of either BNT162b2 or mRNA1273 vaccine, then tested 2-4 months after dose 2. In those participants who informed us that they had decided to get a third vaccine dose, NAb levels were measured within 2 weeks of receiving dose 3 of either BNT162b2 or mRNA1273 and then measured again 2 to 4 weeks after dose 3.

### Patient and Public Involvement

Upon recruitment of participants into the study, we educate them on the difference between any COVID-19 antibody test and a test that semi-quantitatively measures their levels of NAbs(14). This educational component of the recruitment process has resulted in an overwhelmingly positive response by participants to have their levels of NAbs monitored monthly (subsequent study) and recruitment of their friends and relatives into the study. It seems that people want to know how well the vaccine worked for them and how long NAb levels will last. We explain to participants that no study has yet connected NAb levels with protection from disease.

### Determining NAb Levels for a Rapid Test

To perform the test, 10ul whole blood from a finger-stick was added to the LFA cassette sample port followed by 60µl (2 drops) of chase buffer. After 10 minutes, densities of both test and control lines were electronically recorded using an iDetekt RDS-2500 density reader. The test leverages the interaction between RBD-conjugated green-gold nanoshells (Nanocomposix) that bind ACE2 at the test line when RBD-neutralising antibodies (RBD-NAbs) are absent or low. Test line density is inversely proportional to RBD-NAbs present within the sample. As a semi-quantitative test, the results of the LFA can be interpreted using a scorecard or a lateral flow reader.

### Focus Reduction Neutralisation Test (FRNT)

To support the application of the rapid test to measure NAb levels to SARS-CoV-2, we correlated LFA test line densities with IC_50_ values obtained in a Focus Reduction Neutralisation Test (FRNT) from 38 serum samples. Test performance was evaluated using a correlation regression analysis of IC_50_ values and LFA line densities to obtain the equation, Y=-0.7698*X +24.14 when X=log_2_ IC_50_ as shown in Supplementary Figure S2 (**Figure S1**).

### Statistical Analyses

Levene’s test was used to assess homoscedasticity between groups prior to significance testing (IBM SPSS Statistics for Macintosh, Version 26.0; Armonk, NY). To account for unequal variances resulting from unequal sample sizes, Welch’s *t*-test with Benjamini-Hochberg false discovery rate (FDR) correction was performed using Microsoft Excel (Version 16.55; Redmond, WA) to evaluate significant differences in mean neutralisation between BNT162b2 (*n* = 180) and mRNA-1273 (*n* = 89) 2-4 weeks post-2^nd^ dose. Cohen’s *d* was calculated using Microsoft Excel. Post-hoc power analysis was computed using G*Power 3.1 software(15).

## RESULTS

### Correlation of Test Line Densities to Serum IC_50_ values

To support the application of the LFA to measure NAb levels to SARS-CoV-2, we previously reported(14) correlation of LFA test line densities with IC_50_ values obtained in a Focus Reduction Neutralisation Test (FRNT) and demonstrated that the rapid test can accurately and semi-quantitatively measure levels of NAbs directed against SARS-CoV-2. Serum samples with strong neutralising activity demonstrate low line densities while sera with weak neutralising activity demonstrate strong line intensities.

Armed with IC_50_ values, LFA densities and neutralising serum titers from the FRNT, we calculated % neutralisation as: 1-(Test Line Density/Limit of Detection)*100%. **Table 1** shows percent neutralisation ranges that correlate to serum titers, FRNT_50_ values and test line densities. Percent neutralisation was used throughout the study to measure NAb levels in study participants. **Supplementary Figure S2** shows actual LFA tests with density values and corresponding IC_50_s, NAb titers, and percent neutralisation.

**Table 1.**
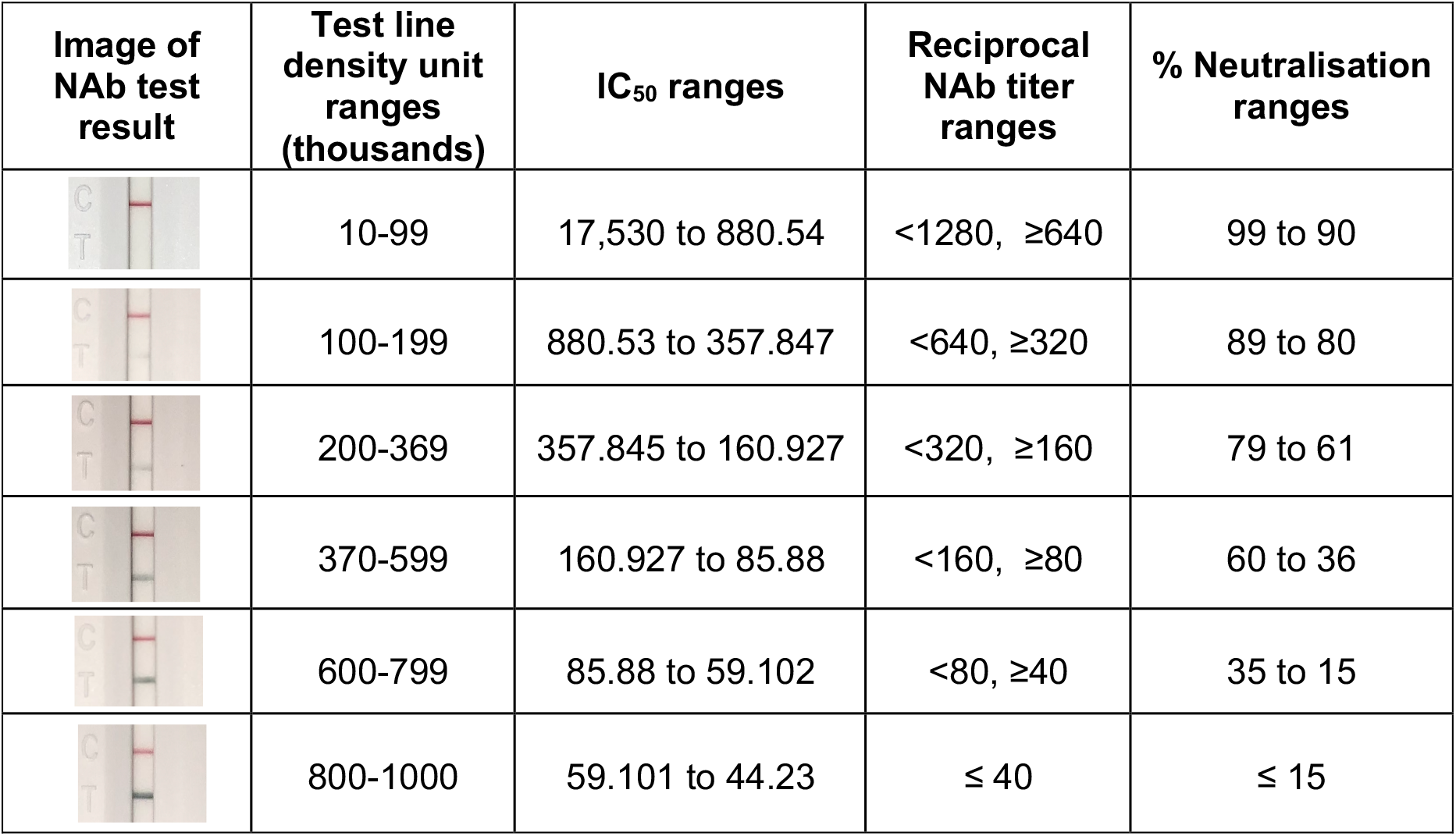
Comparison of LFA Density Units to IC_50_, NAb Titer, and Percent Neutralisation. Correlation of IC_50_ values from a Focus Reduction Neutralisation Test (FRNT) using authentic SARS-CoV-2 with serum titers using 17 PCR-confirmed samples with IC_50_ values <125, six samples with IC_50_ values ≥125 and <250, nine samples with IC_50_ values ≥250 and <500, four samples with IC_50_ values ≥500 and <1000 and three samples with IC_50_ values ≥1000. Reciprocal NAb titers were derived using the highest dilution that did not exceed each serum IC_50_ value. Percent neutralisation was calculated using the following formula: 1-(Test sample line density/Limit of Detection)*100% where LoD for non-neutralising sera for the rapid test was 942,481. Limit of detection (LoD) was calculated based on the method of Armbruster and Pry (16), using a convalescent serum sample containing the lowest detectable concentration of analyte (neutralising antibodies) still distinguishable from a blank. Due to the competitive format of the LFA, the operand was changed to reflect subtraction from limit of blank (LoB) rather than addition: LoD= limit of blank (LoB) – (1.65* SD_low conc sample_): LoD=1,047,382- (1.65 * 63,769)= 942,481 Test Line Density Units. A lower LoD was not applicable, as polyclonal antisera was used in this study, rather than an individual Mab. Alternatively, the average line density observed for the top 10 donors who demonstrated the strongest ability to prevent RBD from binding to ACE2 was 20,706.

| Image of NAb test result | Test line density unit ranges (thousands) | $IC_{50}$ ranges | Reciprocal NAb titer ranges | % Neutralisation ranges |
| --- | --- | --- | --- | --- |
|  | 10-99 | 17,530 to 880.54 | <1280, ≥640 | 99 to 90 |
|  | 100-199 | 880.53 to 357.847 | <640, ≥320 | 89 to 80 |
|  | 200-369 | 357.845 to 160.927 | <320, ≥160 | 79 to 61 |
|  | 370-599 | 160.927 to 85.88 | <160, ≥80 | 60 to 36 |
|  | 600-799 | 85.88 to 59.102 | <80, ≥40 | 35 to 15 |
|  | 800-1000 | 59.101 to 44.23 | ≤ 40 | ≤ 15 |

### COVID-19 RNA Vaccine Evaluation of NAb Response

NAb levels were measured in 269 healthy individuals ranging in age from 19 to 80 (Average age = 51; 165 females and 104 males) who received either BNT162b2 (Pfizer) or mRNA1273 (Moderna) vaccines using our semi-quantitative rapid test(14). NAb levels in vaccine recipients were measured at: i) 2-4 weeks after a second vaccine dose, ii) 2-4 months after the second dose, iii) within 1-2 weeks prior to a third dose and iv) 2-4 weeks after a third RNA vaccine dose. Several observations were made during this study. Percent neutralisation ranged from 0% to 99% 2-4 weeks after a second dose (**Figure 2A**). While our results agree with previous findings in which the majority of vaccine recipients demonstrate NAb levels at ≥75% 2-4 weeks after their second dose(12,13), our study also revealed that 25% of vaccine recipients did not neutralise above 50% (Median neutralisation = 21%) within a month after their second dose (**Figure 2A**).

**Figure 2.**
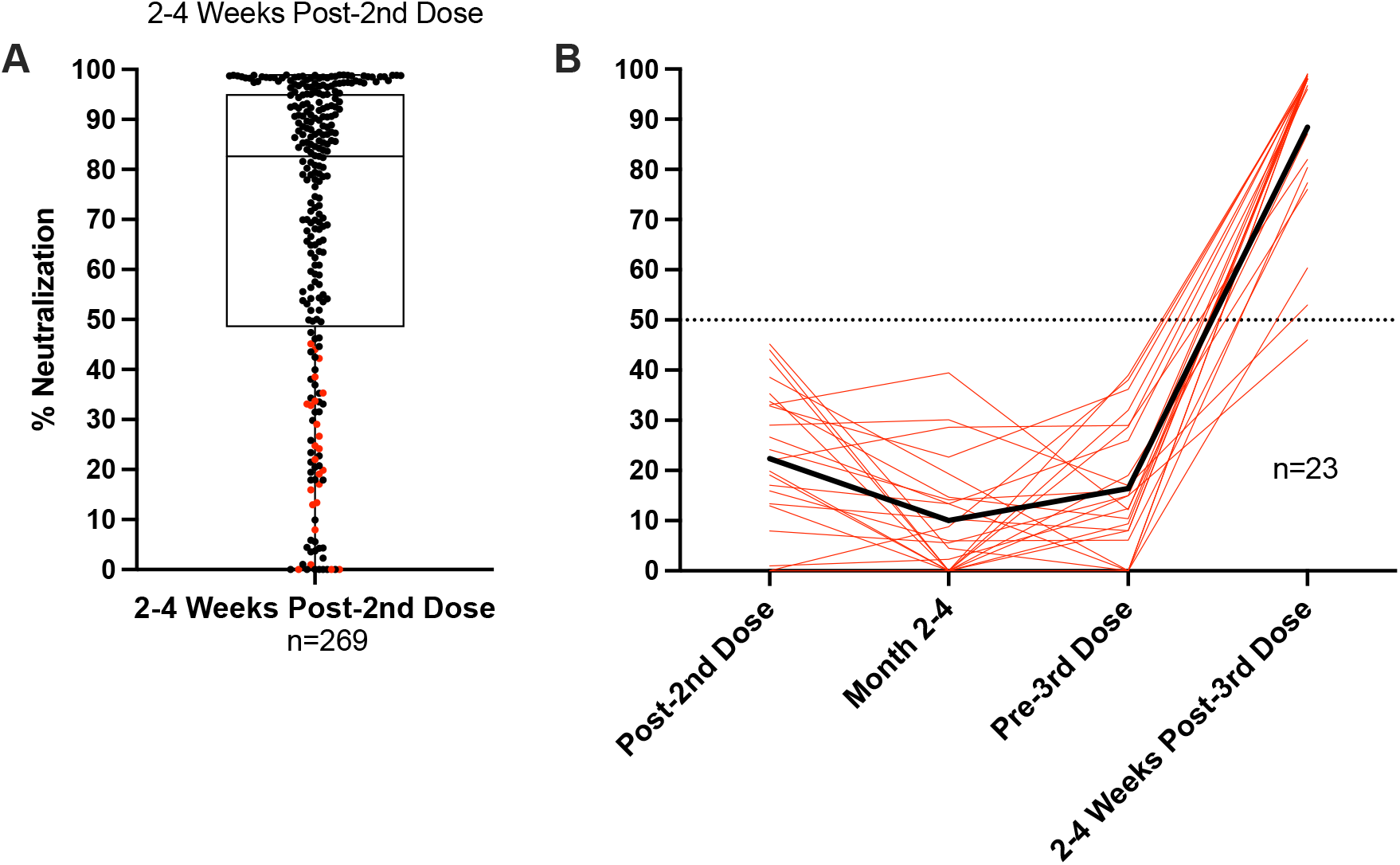
NAb Profile of RNA Vaccine Recipients Pre- and Post-3rd Vaccine Dose. **A) Spectrum of NAb levels 2-4 weeks post 2nd RNA vaccine dose** (180 BNT162b2 combined with 89 mRNA-1273 vaccine recipients=269) ranging from 0% neutralisation to 99% neutralisation. Horizontal line within second and third quartile box denotes median at 83%. Sixty-nine participants in the lower quartile neutralised at <50%. Red dots in lower quartile indicate participants who received 3rd vaccine doses as shown in panel B. **B) Vaccine Poor Responder Third Dose Recipients** Red lines indicate NAb levels of poor responders (<50% neutralisation) at 2-4 weeks post second dose, 1-2 weeks prior to a third vaccine dose and two to four weeks after a third dose of either BNT162b2 or mRNA-1273. Solid black line is the average % Neutralisation of 3rd vaccine dose recipients at each time point. At 2-4 weeks post 3rd dose the average neutralisation was 88%.

Twenty-three VPRs ranging in age from 31 to 79 (10 males, 13 females, average age = 62.5, **Supplementary Table S1**) independently obtained a third dose of either BNT162b2 or mRNA1273 vaccine 1-8 months (average = 5 months) after their second dose. Within a month after their third dose, VPRs showed an average 20-fold increase in NAb levels (range 46%-99%) (**Figure 2B**), suggesting that most VPRs are not permanently poor responders; they are capable of generating high NAb levels with an additional vaccine dose.

Separating VPRs in Figure 2A into mRNA1273 and BNT162b2 vaccine recipients unexpectedly revealed that 14% of mRNA1273 recipients were VPRs, while 31% of BNT162b2 recipients were VPRs. Only one of twelve mRNA1273 VPRs chose to receive a third dose of vaccine. In contrast, 23 of 58 BNT162b2 VPRs chose to receive a third dose of either vaccine (see **Supplemental Table S1)** as shown in **Supplementary Figure S3**. Statistically, Levene’s test indicated heteroscedasticity (*p* < 0.001), while Welch’s *t*-test showed significant differences in mean neutralisation between groups 2-4 weeks post-2^nd^ dose (*q* < 0.001) with medium effect (*d* = 0.537) and observed power nearing unity (1 – β = 0.981).

## DISCUSSION

Some considerations about our findings include the following. We were surprised to observe that 67/269 (25%) of participants in our study did not demonstrate neutralisation >50%. It is not known if poor NAb responders are at increased risk of infection. However, anti-viral T cells and antibodies that mediate ADCC are also important components of immunity and prevent disease once a host is infected. Although 50% neutralisation corresponds to titers <1:160, it is not known if titers of 1:80, for example, would protect an individual from infection. Likewise, it is not known if individuals with highly neutralising antibodies corresponding to titers of ≥1:320 would not be protected from infection. However, some models and reports have predicted that NAb levels can serve as a correlate of protection(6,17).

The debate about whether a vaccinated individual can transmit virus depends in part on their levels of neutralising antibodies. NAbs prevent infection and are used therapeutically to treat COVID-19 patients(9). T cells are crucially important for eliminating infected cells(18–20), but if anti-viral T cells are engaged, the host is already infected. As NAb levels decrease with time after vaccination, there is an increased likelihood that exposure to SARS-CoV-2 could lead to infection which could potentially lead to transmission(21). This may be an important point since a significant portion of the population has not been vaccinated and could be infected by a vaccinated individual whose NAb levels are low, such that they do not prevent infection and asymptomatically shed virus just prior to reactivation of immune memory.

Twenty-five percent of total participants (n=269) in our study did not generate NAb levels stronger than 50% after a 2-dose regimen. These VPRs ranged in age from 19 to 80 with an average age of 57, median age of 60 (n=67). The age range of non-VPRs was 20 to 80 with an average age of 50, median age of 51 (n=202). Further studies could be performed to determine the relationship of age and poor response to COVID-19 vaccination. Our data suggest that COVID-19 vaccine strategies might follow at least a 3-dose regimen to keep peripheral NAb levels high, limiting infection, asymptomatic viral replication, and potential transmission. It also suggests that NAb levels in vaccine recipients could be evaluated with a rapid test on an individual basis to indicate when an additional dose might be indicated.

Although healthcare policy may recommend that a population should receive a third COVID-19 vaccination at a particular time point, an inexpensive rapid test could provide personalised NAb levels on an individual basis to indicate who might or *might not require* a third dose. Not only would this conserve vaccine, but vaccinating individuals who already have elevated levels of NAbs may not provide benefit since spike protein could be cleared by circulating NAbs as fast as it is made by cells.

Previous reports indicate that NAb levels decline much more rapidly than protection from hospitalisation and disease,(12,22) but that does not account for vaccine recipients who never generated high levels of NAbs after two doses. Moreover, it is possible that VPRs could be a source of breakthrough infections. Although it is not known what levels of NAbs protect from infection or disease, many vaccine recipients in high-risk professions may wish to keep peripheral NAb levels high, limiting infection, asymptomatic viral replication, and potential transmission.

### Comparison with other studies

Although vaccine durability studies indicate an average neutralisation geometric mean titer (GMT) of ≥ 320 during the peak period after 2^nd^ dose(10,23), the distribution among individual serum samples obtained during the observed peak neutralisation period (4 to 30 days post-2^nd^ dose) varies greatly(23). It is unclear what percentage of a population falls below a given GMT or IC_50_ during the peak neutralisation period following 2^nd^ dose. Our study supports other findings that majority of healthy individuals generate a NAb response ≥ 75% neutralisation (IC50 ≥160 and <320). However, we highlight a VPR population that, despite healthy status at the time of vaccination, fail to mount a NAb response >50% (IC50 <160) after two doses.

Poor NAb titers have been reported in special populations such as patients with ongoing cancer therapies(24), solid organ transplant patients(25–27), and individuals on systemic immunosuppressive regimens for various immune-mediate inflammatory diseases(28). However, current literature is lacking regarding protective antibody responses to COVID-19 in a healthy population. Finally, it is not unprecedented in other vaccine settings such as influenza to observe poor or non-neutralising responses in healthy individuals(29,30). Due to the urgency to develop vaccines to slow the COVID-19 pandemic, we are still learning the parameters of RNA vaccine dose, frequency, timing and durability in the human population.

### Limitations of the study

This study has several limitations. First, it is still unknown what levels of neutralising antibodies correlate with protection against infection and potential disease. It is possible, but unlikely, that NAb levels as low as 20% could protect against infection(6). Second, although the N-terminal domain of spike protein also been shown to neutralise SARS-CoV-2, it is currently characterised as a minor component of neutralising antibodies(7,31) and our test does not detect them. Although we measured NAb levels for twice as many BNT162b2 vaccine recipients as mRNA-1273 recipients, we examined homogeneity of variance using Levene’s test and, upon confirming unequal variances, assumed Welch’s *t*-test as a conservative and robust alternative to parametric comparisons of means. Importantly, potential for type I error was mitigated using FDR-adjustment of calculated significance, and Cohen’s *d* showed appreciable effect size between groups. Moreover, post-hoc power analysis showed exceptional sensitivity and low chance of type II error, further supporting the significantly lower percent neutralisation observed in Pfizer recipients 2-4 weeks post-2^nd^ dose.

## Conclusions

In conclusion, our findings suggest that 14% of mRNA1273 and 31% of BNT162b2 two-dose vaccine recipients ranging in age from 19 to 80 with an average age of 57 (median age of 60) may not have generated levels of NAbs ≥50% and that additional COVID-19 vaccine doses might be indicated for these individuals. Longitudinal studies are ongoing to determine if high NAb levels in recipients of a third vaccine dose are more durable than NAb levels after two doses.

### What is already known on this topic

- NAb levels and T cell immunity decline after vaccination, but that doesn’t account for those vaccine recipients who did not make strong NAb responses after vaccine dose 2.
- Mixing and matching vaccines is not harmful.
- Most healthy individuals respond to 2 vaccine doses with high NAb titers

### What this study adds

- Vaccine recipients who did not generate strong NAb responses to the first 2 doses require a third dose to generate high levels of NAbs.
- 25% of BNT162b2 vaccine recipients may not know that they do not neutralise SARS-CoV-2 more strongly than 50%.
- Strong NAb responses to a third dose of RNA vaccine appear to be independent of manufacturer.

## Data Availability

All de-identified raw data corresponding to results shown as part of the study demonstrated in this manuscript can be made available upon request to corresponding author.

## Ethics Statement

### Ethical approval

All data generated in this study used finger-stick peripheral blood collected under an Arizona State University institutional review board (IRB) approved protocol #0601000548. Subjects were assigned a vaccine study de-identification number (VAC-ID) at the time of enrollment and all subsequent collections were conducted in compliance with the Collaborative Institutional Training Program (CITI) Human Subjects Research (HSR) regulatory guidance.

## Acknowledgements

We thank Arizona State University School of Life Sciences and Mayo Clinic Arizona Collaborative Research facilities for their providing the laboratory space and time needed to conduct this study at multiple testing location sites. We thank Empowered Diagnostics for providing the test strips for this study.

## Footnotes

### Contributors

DFL and AJR contributed equally to this work as joint first authors, writing the manuscript, generating and editing figures. SS, MGM, and AS-N contributed to development of the NAb LFA and supplied test materials. AJR and MAK conducted data analysis and collection. MAK, JV, DM, JF, BE helped with data collection and recording. EJK provided insight to study design. PZ contributed to statistical analysis.

### Funding

This study was funded in part by Sapphire/AXIM Biotechnologies, Inc. (San Diego, CA). The funder had no role in the study design, collection, analysis, and interpretation of data.

### Competing Interests

All authors have completed the *Unified Competing Interest form* and declare: no support from any organisation for the submitted work except DFL and SS are co-founders of Sapphire, the research division of AXIM Biotechnologies. SS, MGM, and AS-N are employed by AXIM. All other authors have no competing interests and no financial relationships with any organisations that might have an interest in the submitted work in the previous three years, no other relationships or activities that could appear to have influenced the submitted work.

## SUPPLEMENTARY MATERIALS

**Supplementary Figure S1.**
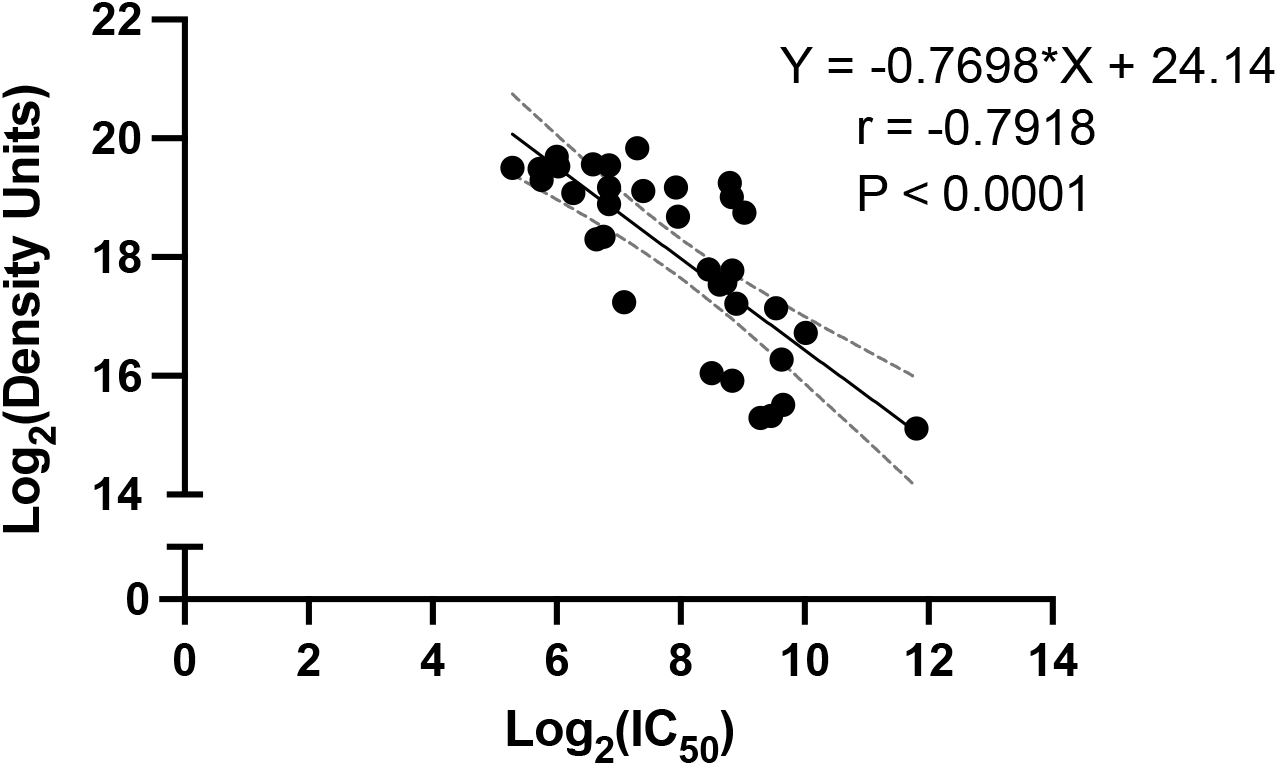
Regression analysis of LFA density values and microneutralisation IC_50_ values. Comparison between LFA density units and IC_50_ values obtained using convalescent sera isolated from 36 recovered COVID-19 patients. Neutralisation was tested on LFA using 6 µL serum and a dilution series was performed by authentic SARS-CoV-2 FRNT assay to obtain IC_50_ values. To calculate IC_50_, data were analyzed in GraphPad Prism 9.0 using methods described by Ferrara and Temperton. Density values and IC_50_ values were Log_2_-transformed and analyzed using a simple linear regression and nonparametric Spearman correlation with two-tailed P value and a 95% confidence interval (CI). Regression analysis with 95% CI boundaries is indicated in solid black and grey dotted lines. Spearman’s rho and two-tailed P value are labeled.

**Supplementary Table S1.**
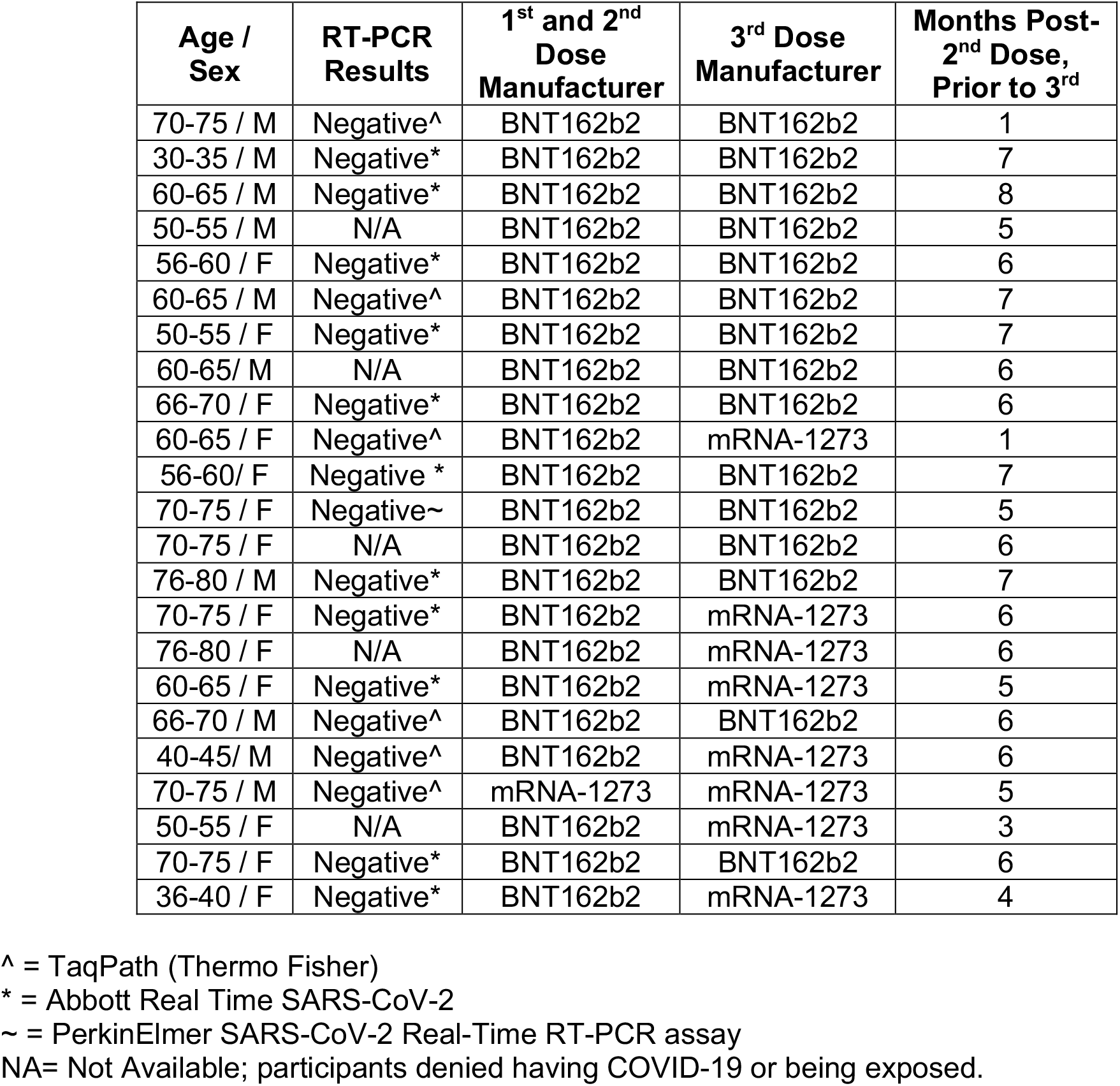
Demographic Data of Poor Responders with a 3^rd^ RNA Vaccine Dose. Demographic information for 23 study participants who received a 3^rd^ RNA Vaccine Dose. Twenty-two individuals received two doses of BNT162b2 and one individual received two mRNA-1273 doses initially. 15 of the 22 individuals that were originally vaccinated with BNT162b2 obtained a 3^rd^ dose of BNT162b2, and 8 received mRNA-1273 (100µg) as their 3^rd^ dose. One individual originally vaccinated with mRNA-1273 received a 3^rd^, 100µg dose of mRNA-1273. All participants had either confirmed negative RT-PCR results or no known history of infection prior to enrollment. RT-PCR platform indicated using symbols defined below Table S1. Age ranges are provided to protect the identities of the individuals in the study.

| Age / Sex | RT-PCR Results | 1<sup>st</sup> and 2<sup>nd</sup> Dose Manufacturer | 3<sup>rd</sup> Dose Manufacturer | Months Post-2<sup>nd</sup> Dose, Prior to 3<sup>rd</sup> |
| --- | --- | --- | --- | --- |
| 70-75 / M | Negative<sup>^</sup> | BNT162b2 | BNT162b2 | 1 |
| 30-35 / M | Negative<sup>*</sup> | BNT162b2 | BNT162b2 | 7 |
| 60-65 / M | Negative<sup>*</sup> | BNT162b2 | BNT162b2 | 8 |
| 50-55 / M | N/A | BNT162b2 | BNT162b2 | 5 |
| 56-60 / F | Negative<sup>*</sup> | BNT162b2 | BNT162b2 | 6 |
| 60-65 / M | Negative<sup>^</sup> | BNT162b2 | BNT162b2 | 7 |
| 50-55 / F | Negative<sup>*</sup> | BNT162b2 | BNT162b2 | 7 |
| 60-65 / M | N/A | BNT162b2 | BNT162b2 | 6 |
| 66-70 / F | Negative<sup>*</sup> | BNT162b2 | BNT162b2 | 6 |
| 60-65 / F | Negative<sup>^</sup> | BNT162b2 | mRNA-1273 | 1 |
| 56-60 / F | Negative<sup>*</sup> | BNT162b2 | BNT162b2 | 7 |
| 70-75 / F | Negative<sup>~</sup> | BNT162b2 | BNT162b2 | 5 |
| 70-75 / F | N/A | BNT162b2 | BNT162b2 | 6 |
| 76-80 / M | Negative<sup>*</sup> | BNT162b2 | BNT162b2 | 7 |
| 70-75 / F | Negative<sup>*</sup> | BNT162b2 | mRNA-1273 | 6 |
| 76-80 / F | N/A | BNT162b2 | mRNA-1273 | 6 |
| 60-65 / F | Negative<sup>*</sup> | BNT162b2 | mRNA-1273 | 5 |
| 66-70 / M | Negative<sup>^</sup> | BNT162b2 | BNT162b2 | 6 |
| 40-45 / M | Negative<sup>^</sup> | BNT162b2 | mRNA-1273 | 6 |
| 70-75 / M | Negative<sup>^</sup> | mRNA-1273 | mRNA-1273 | 5 |
| 50-55 / F | N/A | BNT162b2 | mRNA-1273 | 3 |
| 70-75 / F | Negative<sup>*</sup> | BNT162b2 | BNT162b2 | 6 |
| 36-40 / F | Negative<sup>*</sup> | BNT162b2 | mRNA-1273 | 4 |

<sup>^</sup> = TaqPath (Thermo Fisher)
<sup>\*</sup> = Abbott Real Time SARS-CoV-2
<sup>~</sup> = PerkinElmer SARS-CoV-2 Real-Time RT-PCR assay
NA= Not Available; participants denied having COVID-19 or being exposed.

**Supplementary Figure S2.**
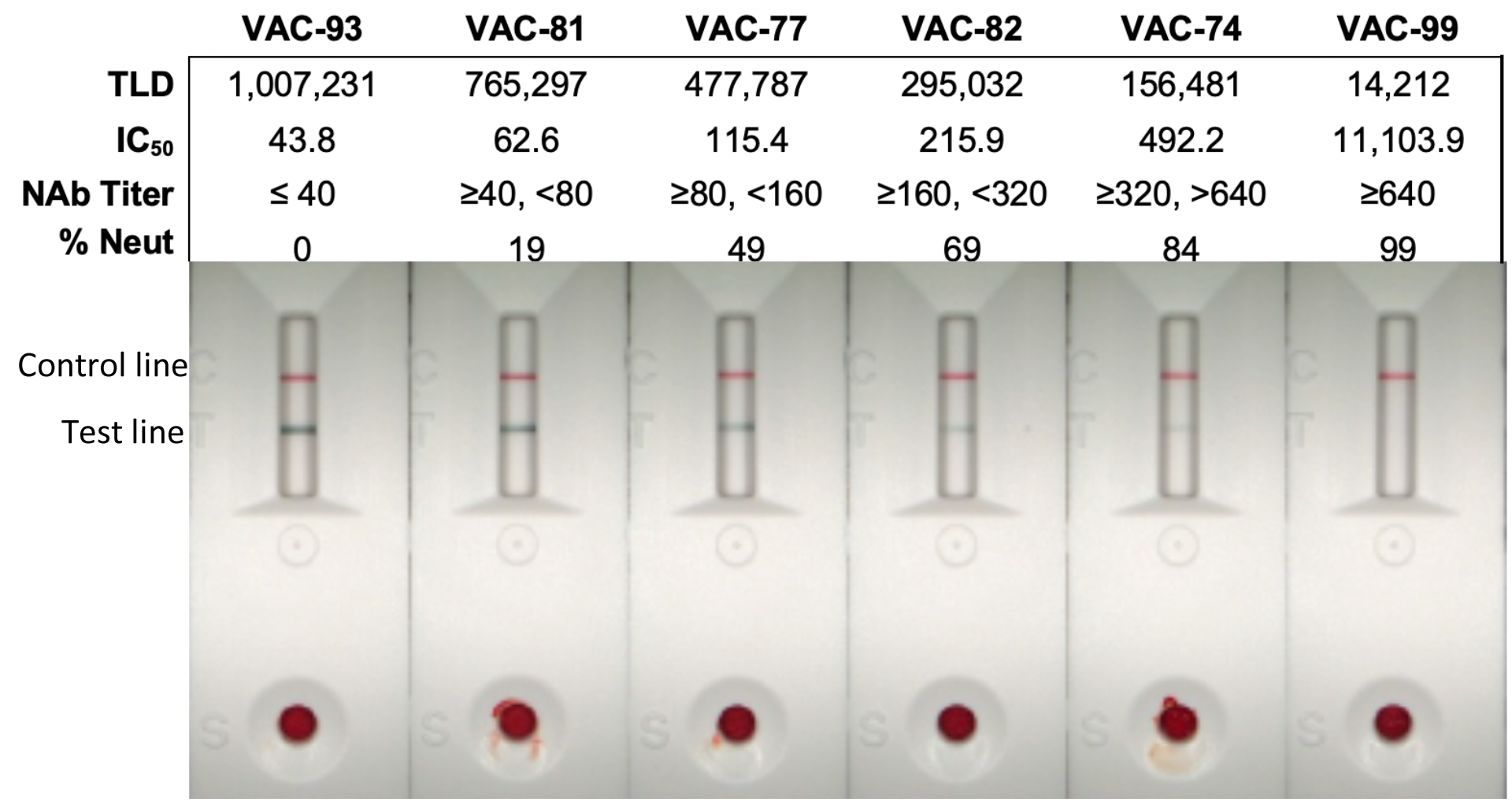
NAb LFA Tests with Density Values, IC50s, NAb Titers, and Percent Neutralisation. LFA densitometer images with corresponding Test Line Density (TLD), IC_50_ value, reciprocal NAb titer, and percent neutralisation. TLD is reported as unmodified, discrete values. IC_50_ values are derived from the regression equation shown in Supplementary **Figure S1**. Reciprocal NAb titer is derived from IC_50_, such that titer is equal to the last dilution factor (DF) (DF =20*2^-n^) for which IC50 is ≥ the lower threshold range for a given titer. For example, IC_50_=43.8 is classified as an NAb titer of 40. Further, a hypothetical IC_50_=158 would classify as NAb titer ≥80 and <160. Percent neutralisation was calculated using the equation: 1-(TLD/LoD) when LoD=942,481 as described in **Table 1** legend.

**Supplementary Figure S3.**
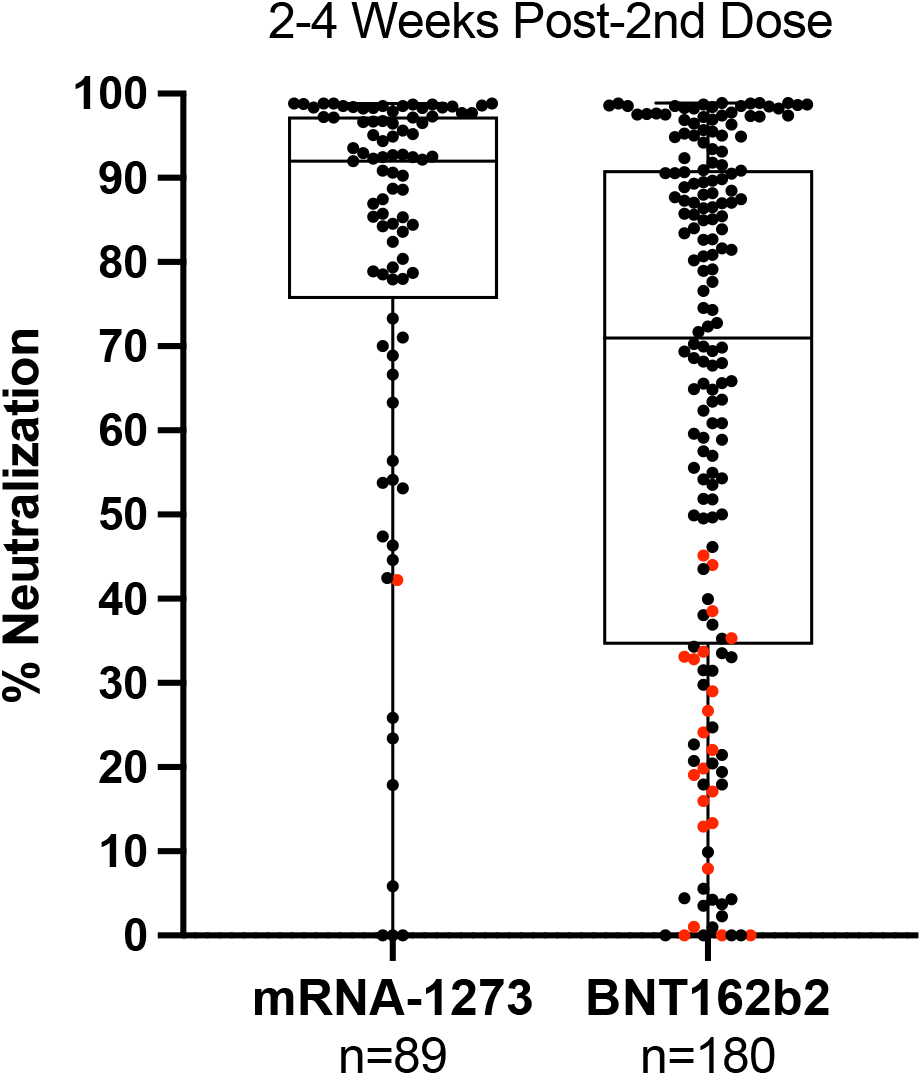
Comparison of NAbs 2-4 weeks after 2^nd^ Dose of BNT162b2 or mRNA1273. Comparison of NAbs post-2^nd^ dose according to vaccine received (mRNA1273 or BNT162b2) ranging from 0% to 99% neutralisation. Data shown as box and whisker plots with black vertical lines that denote upper and lower extremes, and horizontal lines that denote upper and lower quartiles with median at the midline. Median neutralisation of mRNA1273 (n=89) and BNT162b2 (n=180) is 92% and 71%, respectively. Mean neutralisation for mRNA1273 and BNT162b2 groups is 80% and 63%, respectively. Red dots indicate VPRs that received a 3^rd^ vaccine dose as shown in **Figure 2B** and described in Supplementary **Table S1**.

## Notes

### Funding Statement

This study was partially funded by Axim Biotech, San Diego, CA

### Author Declarations

Arizona State University Internal Review Board

